# Why do per capita COVID-19 Case Rates Differ Between U.S. States?

**DOI:** 10.1101/2020.10.16.20213892

**Authors:** Lloyd Chambless

## Abstract

**Background:** The popular press has explored the differences among U.S. states in rates of COVID-19 cases, mostly focusing on political party differences, and often mentioning that political party differences in health outcomes are confounded by demographic and socio-economic differences between Democratic areas and Republican areas. The purpose of this paper is to present a thorough analysis of these issues.

**Design and Methods:** State-specific COVID-19 cases per 100,000 people was the main outcome studied, with explanatory variables from Bureau of Census surveys, including percentages of the state population that were Hispanic, black, below poverty level, had at least a bachelor’s degree, or were uninsured, along with median age, median income, population density, and degree of urbanization. We also included political party in power as an explanatory variable in multiple linear regression. The units of analysis in this study are the 50 U.S. states.

**Results:** All explanatory variables were at least marginally statistically significantly associated with case rate in univariate regression analysis, except for population density and urbanization. All the census characteristics were at least marginally associated with party in power in one factor analysis of variance, except for percentage black. In a forward stepwise procedure in a multivariable model for case rate, percentages of the state population that were Hispanic or black, median age, median income, population density, and (residual) percentage poverty were retained as statistically significant and explained 62% of the variation between states in case rates. In a model with political party in power included, along with any additional variables that notably affected the adjusted association between party in power and case rate, 69% of the variance between states in case rates was explained, and adjusted case rates per 100,000 people were 2155 for states with Democratic governments, 2269 for states with mixed governments, and 2738 for Republican-led states. These estimates are based on data through October 8, 2020.

**Conclusions:** U.S. state-specific demographic and socio-economic variables are strongly associated with the states’ COVID-19 case rates, so must be considered in analysis of variation in case rates between the states. Adjusting for these factors, states with Democrats as the party in power have lower case rates than Republican-led states.

## Introduction

The COVID-19 pandemic of 2020-21 will induce a plethora of publications exploring the variation in outcomes among the countries of the world and among states in the U.S. These outcomes will be economic, social, political, and, above all, health-related. The health-related outcomes include number of COVID-19 cases, number of hospitalized cases, and number of deaths. Indeed, many websites are already reporting the health outcomes in the U.S. by county and by state, and there have been multiple articles in the popular press exploring the differences between states.^1-6^ Most of the articles focus on political party differences between states or counties, and many mention that political party differences in health outcomes are confounded by demographic, geographic, and socio-economic differences between Democratic areas and Republican areas. One article claims that it is these confounding factors that explain the differences between Democratic and Republican areas, though no formal statistical analysis of this issue is offered.^4^ The purpose of this paper is to present the results of such an analysis, in which we quantify the associations between those factors and both COVID-19 and political party, and then quantify the difference between Democratic and Republican areas in health outcome after controlling for these confounding factors. Future studies by many groups will explore the more important issue of which public health measures, such as timing of shutdowns, easing of restrictions, face-mask mandates, crowd-size limitations, and closing of bars, had the greatest effect on COVID-19 outcomes, and how the implementation of these measures relate to partisan differences. Such studies will also need to account for the confounding factors we consider here.

## Methods

### Health Outcome

Our main outcome of interest is confirmed or probable COVID-19 cases per capita. We will use the word “rate” for this outcome, though it is simply a count of cumulative events by state from the beginning of the pandemic in the U.S. through October 8, 2020, divided by the estimated state population in 2018,^7^ and then multiplied by 100,000 to give the “rate” per 100,000 people in the state. The counts of COVID-19 cases by state were taken from a New York Times database.^8^ The following description is condensed from the Times’ description of the database. The Times has compiled this data from state and local governments and health departments. The counts of cases include both laboratory confirmed and probable cases using criteria that were developed by states and the federal government.^9^ Confirmed deaths are individuals who have died and meet the definition for a confirmed Covid-19 case. The Times follows health departments in removing non-Covid-19 deaths among confirmed cases when they have information to unambiguously know the deaths were not due to Covid-19. Probable deaths are deaths where Covid-19 is listed on the death certificate as the cause of death or a significant contributing condition, but where there has been no positive confirmatory laboratory test. Because of the possible association between the level of testing for COVID-19 and the number of cases found, we would have liked to also analyze the rate of COVID-19 hospitalizations by state, but fourteen states do not report this data (The Covid Tracking Project).^10^ We do briefly consider confirmed or probable COVID-19 death rate, again per 100.000 people.^8^

### State Characteristics

Our data come from the U.S. Census Bureau’s 2018 American Community Survey 1-Year or 5-year Estimates, and include percentage of people living in a household with income below the poverty level^11^; median income^12^; percentage of the population living in an urban area;^13^ population density (number of people per square mile);^13^ percentage of the state population that is Hispanic of any race, and the percentage of the population that is African-American and not hispanic;^14^ median age;^15^ percentage of the population above the age of 25 with a bachelor’s degree or higher;^16^ and the percentage of the population aged 19-64 that is uninsured.^17^

### Political status of the states

There are several ways to define this variable. Our primary method is to classify states into three categories according to the party in power: those with governor and both chambers of the state legislature Republican, those with governor and both chambers of the state legislature Democratic, and the others, with mixed government.^18^ Another method of looking at political status is to use the percentage of the vote for the U.S. Congress in 2018 that was Democratic, out of the vote that was either Democratic or Republican^19^. A third way is to use the percentage of the vote for U.S. President in 2016 that was Democratic, out of the vote that was either Democratic or Republican.^20^ Note that it is entirely equivalent to use percentage Republican instead of percentage Democratic. As an alternative, and one that has direct comparison to the party-in-power analysis, instead of using the continuous percentage of vote variables we also divided the states into three groups of the same sizes as the number of Republican, Mixed, and Democratic states, according to the lowest, middle, and highest percentage of Democratic vote, respectively. We did this both for the 2016 presidential vote and the 2018 House vote. A more standard division of the states into three groups according to percentage Demographic vote would be to make the three groups equal in size, which we also considered, with 17, 16, and 17 states in the low, middle, and high groups.

### Time

Since the date of onset of the pandemic varied from state to state, the length of observation period varies, so we will also consider in our models for case rate the date of the first case in each state.^8^ Since case fatality for COVID-19 may be changing over time, we will consider a date variable in our models for death rate that reflects the middle of the pandemic for the state. Our variable is defined as the first date when the cumulative number of deaths in a state has exceeded half the total number of deaths in the period under consideration, which is the median of COVID-19 death dates for the state in the period considered.

### Analysis

The unit of analysis is states, not individuals. We present the graphs of case rate versus each of the state characteristics, giving the best fitted line through the points in each graph, along with the p-value for the association, which indicates the degree of statistical significance (standard univariate least-squares regression). We also present the unadjusted means of case rate by party in power (Democratic, Republican, or Mixed), with the p-value for the differences in these means by party in power (analysis of variance). Some of the differences in case rates between the three government types are possibly related to associations between the state characteristics with both case rate and party in power, so we also present the means of the state characteristics by party in power (analysis of variance). To study the association between party in power and case rate it is important to control for the state characteristics, in order to assess the level of association that is independent of those characteristics. To this end we apply multiple linear regression (analysis of covariance). Our model-building is a forward-building process, in which we add the most important (by p-value) additional state characteristic at each step, as long as a statistically significant variable (p<0.05) can be added. In preparation we consider the correlation between the various state characteristics, and we will see that median income, percentage living in poverty, and percentage with a bachelor’s degree are highly correlated. Thus, we include no more than one at a time of these three highly correlated state characteristics in our models, and in this multiple variable process, median income is selected first among the three in the analysis of case rate. To further consider the other two highly correlated characteristics we replace each by the amount it exceeds its predicted value, as predicted by the other state characteristics, including median income, using linear regression. These differences, called “residuals”, are not correlated with median income nor are they highly correlated with each other, and we include them in our model if the addition is statistically significant. We next add the categorical variables representing party in power to the model, and then add any of the not-yet-included state characteristics that have a notable effect (>10%) on the estimates of adjusted differences between government types, since we want to be certain to include any state characteristic that affects (confounds) the estimates of those differences.

To evaluate whether the date of the onset of COVID-19 in each state affects the conclusions of the analysis, we add this date to the model reached by the steps outlined above.

In a secondary analysis, instead of classifying states by party in power, we study the association between case rate and the Democratic percentage of the 2016 presidential vote or the Democratic percentage of the 2018 House vote. We first present the univariate graphs for these potential associations. We then replace the party-in-power variables in our final model with either percentage of state vote in the 2016 presidential election vote that was Democratic or the percentage of the 2018 federal House vote that was Democratic.

The analysis of death rate is similar to that for case rate, except we use as a date variable the median date of deaths in a state in the period considered.

We also confirm the results of the analyses done with ordinary multiple linear regression using a method well-suited to count data, namely a multiplicative negative binomial regression model using SAS Proc Genmod. All analysis was done with SAS 9.4. Confidence intervals and p-values must be interpreted with caution due to the limited sample size – fifty states.

## Results

There were 15 states with governor and both houses of the legislature Democratic, 21 states similarly Republican, and 14 other states with mixed government. The correlation coefficients among the state characteristics are less than 0.64 except: between median income and both of percentage with bachelor’s degree and percentage living in poverty, for which both correlation coefficients are 0.84 in absolute value, and between percentage in poverty and percentage with bachelor’s degree, for which the correlation coefficient is −0.75.

In the graphs of case rate versus each of the state characteristics (Figure 1), the observed rates (circles) are shown for each state along with the predicted rate (line) for each characteristic, from univariate regression. The p-value given is a test of whether the slope of the association differs from zero, with p < 0.05 usually accepted as statistically significant. Most associations are statistically significant, except for the association with population density and the percentage urban. Table 1 gives the unadjusted means of the state characteristics by government type, along with a p-value for the test of differences between the means by government type. The differences are all statistically significant, or close, except for percentage black. The Democratic states have a greater population density, percentage Hispanic, percentage urban, and median income than the other states. Republican states have a lower median age and percentage with a bachelor’s degree than the other states and a higher percentage uninsured and percentage in poverty. The mixed government states generally have mean state characteristics between those of the Republican and Democratic led states. The unadjusted mean case rates per 100,000 persons are 2850, 1873, and 1870 for Republican, Mixed, and Democratic states, respectively (p < 0.001 for differences between the three).

**Table 1.**
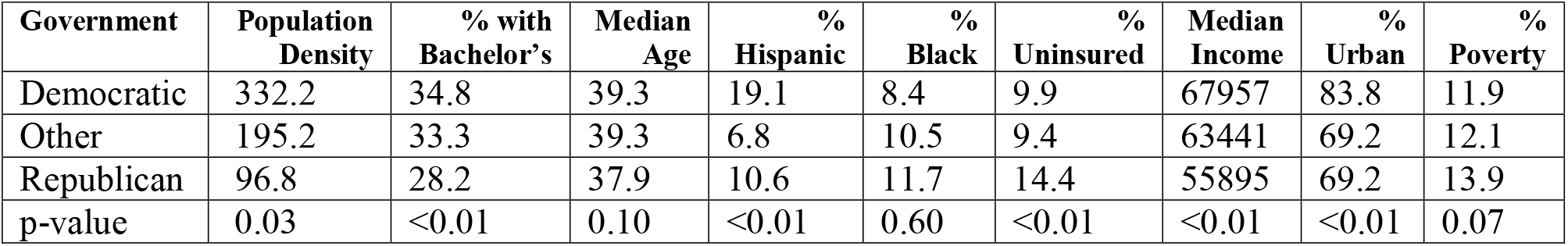
Unadjusted Means of Population Characteristics by State Government Type, with p-value for Test of Differences

| Government | Population Density | % with Bachelor's | Median Age | % Hispanic | % Black | % Uninsured | Median Income | % Urban | % Poverty |
| --- | --- | --- | --- | --- | --- | --- | --- | --- | --- |
| Democratic | 332.2 | 34.8 | 39.3 | 19.1 | 8.4 | 9.9 | 67957 | 83.8 | 11.9 |
| Other | 195.2 | 33.3 | 39.3 | 6.8 | 10.5 | 9.4 | 63441 | 69.2 | 12.1 |
| Republican | 96.8 | 28.2 | 37.9 | 10.6 | 11.7 | 14.4 | 55895 | 69.2 | 13.9 |
| p-value | 0.03 | <0.01 | 0.10 | <0.01 | 0.60 | <0.01 | <0.01 | <0.01 | 0.07 |

**Figure 1:**
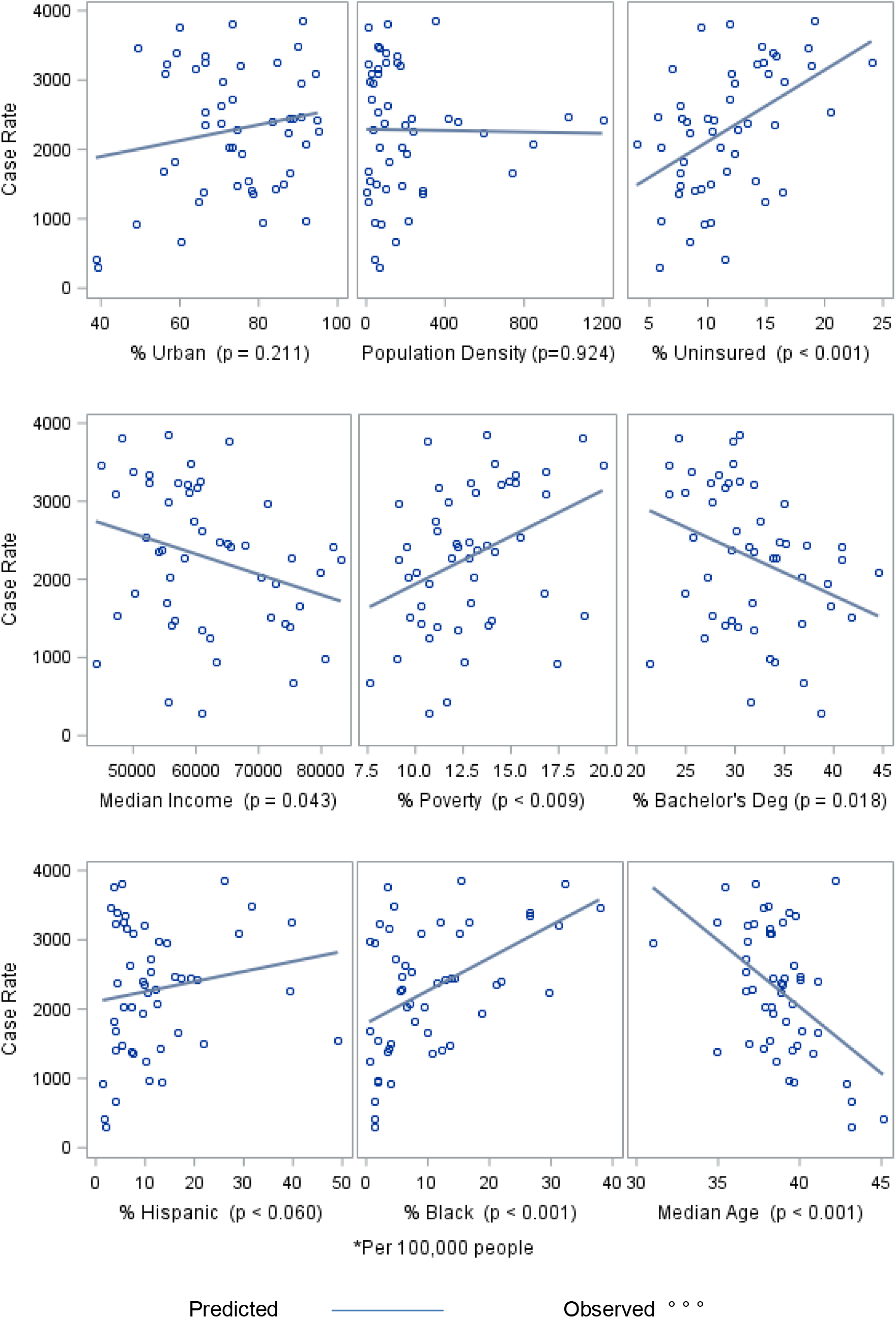
Observed and Predicted Case Rate* versus State Characteristics in Univariate Analysis

Next, we turn to modeling case rates in terms of multiple state characteristics. In building the linear model to accomplish this task, we retained the race/ethnicity, median age, median income, population density, and residual percentage poverty (the excess of observed percentage of poverty beyond that predicted by median income and the other state characteristics) variables, in that order of importance. This model explained 62% of the variance in case rates among the states (R-squared = 0.62). The other state characteristics, including the excess of percentage with bachelor’s degree beyond that predicted by median income and the other state characteristics, had no additional statistically significant association with case rate.

It is instructive to look further at our linear model at this point, before adding the party-in-power variables to the model. In the graph of the observed case rates versus those predicted by the state demographic and socio-economic characteristics (without consideration of party in power) (Figure 2), the diagonal line is the line of equality, so that states above the line have a greater COVID-19 rate than that predicted by state characteristics – the higher above the line, the greater the absolute excess in observed case rate compared to predicted. Similarly, those states below the line have a lower COVID-19 rate than that predicted. The states are labeled for the color of the political party governing, blue for Democrats, red for Republicans, and black for mixed governments. The preponderance of points far above the line are red.

**FIGURE 2:**
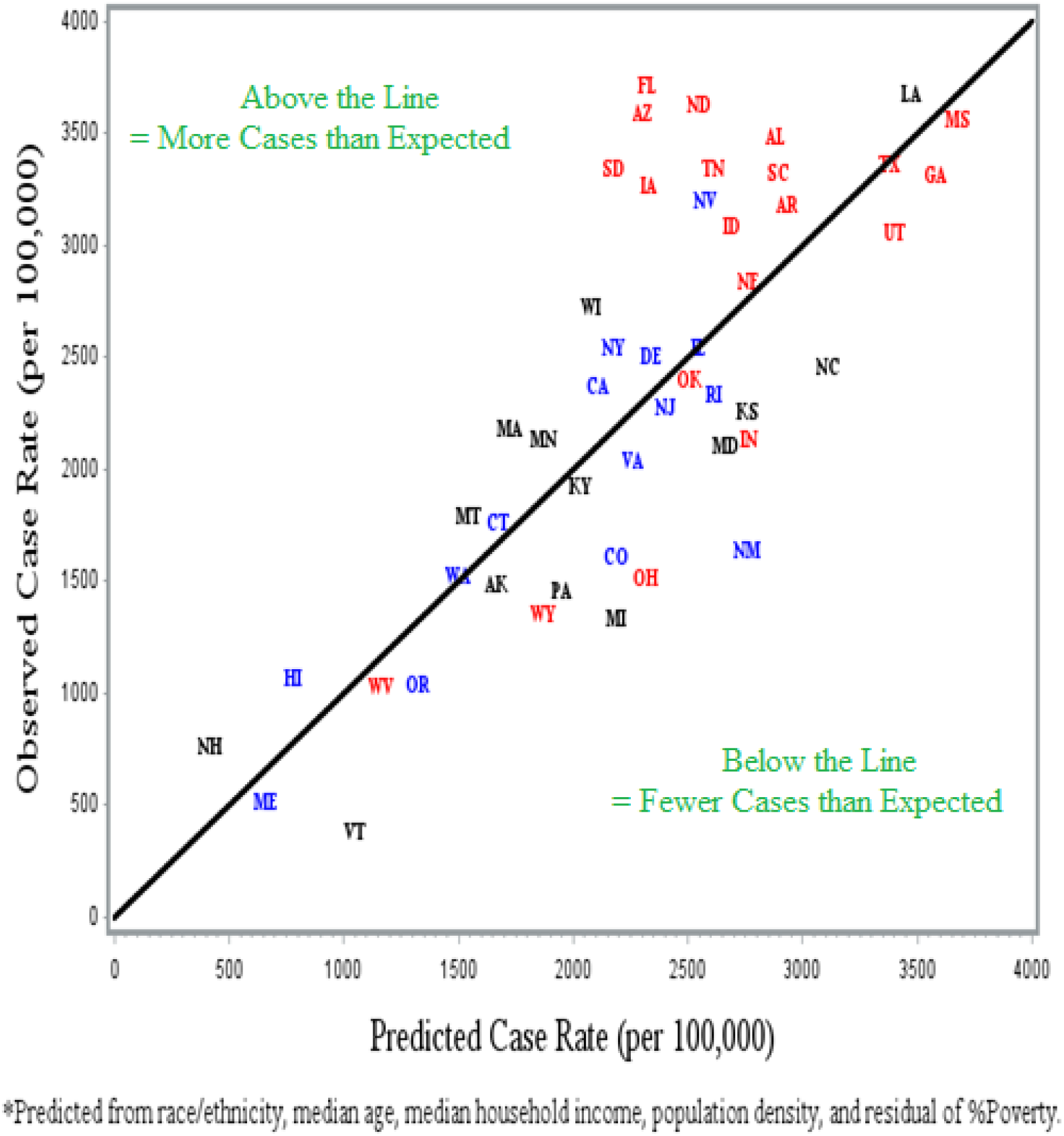
Observed versus Predicted* Cumulative Covid 19 Cases per 100,000 Papulation. Patty in Power: Democratic=Blue, Republican = Red, Other = Black

To estimate the mean differences between the three types of government, adjusted for the state characteristics, we add party-in-power variables to the predictive model above. Before finalizing this model, we first explore whether any of the state characteristics not in the predictive model would nevertheless affect our estimates of the adjusted differences between states. Neither of percentage urban nor percentage uninsured did so, nor are they statistically significant, so they are not considered further. On the other hand, ‘excess of percentage with bachelor’s degree beyond that predicted by median income and the other state characteristics’ notably (more than 10%) affected our adjusted differences between states, so we retained this variable in our final model. This final model explains 69% of the variation between the states in case rates, and estimates of the effects of each variable in the model, adjusted for the other variables, are given in Table 2. To express the effect of a continuous state characteristic, two levels of the characteristic are compared. A somewhat arbitrary but frequently used comparison is adopted here, between the 75^th^ percentile and the 25^th^ percentile of the variable. Those percentiles are given along with the difference in predicted case rate at the 75^th^ percentile minus that at the 25^th^. We also include the 95% confidence intervals for these differences. Table 3 indicates, for example, that states at the 75^th^ percentile of percentage Black (14.3%) have a predicted case rate that is 389 per 100,000 higher than states at the 25^th^ percentile (3.4%), with a 95% confidence estimate of the difference being (177, 602) per 100,000. Percentage Black and median age have statistically significant effects on case rate, adjusted for the other variables in the model. The table further gives the estimates that Mixed government states have a 469 cases per 100,000 lower case rate than do Republican-led states (p = 0.051) and that Democratic-led states have a 583 cases per 100,000 lower case rate than Republican states (p = 0.041). An alternative presentation of this result is in terms of the predicted rates for average values of the state characteristics: predicted case rates per 100,000 people were 2155 for Democratic states, 2269 for mixed government states, and 2738 for Republican states. Thus, Democratic states and mixed government states had case rates 79% and 83%, respectively, of the Republican states. The results from the negative binomial regression model were similar, at 73% (p = 0.042) and 79% (p = 0.121), respectively.

**Table 2.** Predicted Increase in Case Rate* from 25^th^ to 75^th^ Percentile of State Characteristic, and Difference in Case Rate by Party in Power Compared to Republicans in Power

|  |  |  |  |
| --- | --- | --- | --- |
| **Characteristic** | **25<sup>th</sup> Percentile** | **75<sup>th</sup> Percentile** | **Predicted Increase in Case Rate from 2nd to 3<sup>rd</sup> Column** |
| % Hispanic | 5.1 | 14.2 | 135 (-39, 310) |
| % Black | 3.4 | 14.3 | 389 (177, 602) |
| Median Age (years) | 37.3 | 39.7 | -417 (-634, -201)) |
| Median Income (\$) | 55248 | 59898 | -104 (-215,7) |
| Population Density (persons/sq. mi.) | 43.1 | 211.8 | 143 (-3, 288) |
| Observed % Bachelor’s Deg minus Predicted % Bachelor’s Degree | -1.5 | 2.0 | -139 (-374, 96) |
| Observed % Poverty minus Predicted % Poverty | -0.76 | 0.43 | -247 (-468, -26) |
| Party in Power | Republican | Democratic | -583 (-1122, -43) |
|  | Republican | Mixed | -469 (-926, -13) |

\*Cases per 100,000 people

It is possible that the results above could be confounded by the date of first case in each state, but when we included that date in the model just presented, there were only negligible changes to the conclusions. In this model, mixed government states have a 455 cases per 100,000 lower case rate than do Republican led governments (p = 0.053) and Democratic governments have a 524 cases per 100,000 lower case rate than Republican states (p = 0.061), and the date variable was not statistically significant (p = 0.097). Thus, to be consistent with the steps we outlined in the methods section for including variables in our model, we will still refer to the model in the previous paragraph as our final model.

In our secondary analysis of percentage Democratic of state vote in the 2016 presidential election or percentage Democratic of state vote in the 2018 U.S. House election, the associations between percentage Democratic and case rate, unadjusted for any state characteristics, were statistically significant for 2016 presidential vote (p = 0.021) but not for 2018 House vote (p = 0.094). (Figure 3). To evaluate the same associations but this time adjusted for the state characteristics, we put in our final party-in-power model one or the other of the two Democratic percentage of vote variables in place of the party-in-power variable. Neither Democratic House vote nor Democratic presidential vote was significantly associated with case rate (p = 0.975 House, 0.107 president) when adjusting for state characteristics, though the results were in the same direction – the higher the percentage of Democratic vote, the lower the predicted case rate. When we instead fit the model with the states divided into three groups of the same size as the three party-in-power groups, in terms of low, middle, and high percentage of Democratic votes in the 2016 presidential election, the high Democratic presidential vote group of states had 894 per 100,000 fewer COVID-19 cases than the low group (p =0.033), and the middle Democratic presidential vote group of states had 490 per 100,000 fewer COVID-19 cases than the low group (0.114). The parallel results for division into three groups of states according to 2018 House vote were that the high Democratic presidential vote group of states had 252 per 100,000 fewer COVID-19 cases than the low group (p =0.358), and middle Democratic House vote group of states had 36 per 100,000 fewer COVID-19 cases than the low group (0.874). Use of three equal groups for percentage vote Democratic gave very similar results to the unequal group analysis just presented, for both presidential and House votes. Doing the analysis simply dividing states into those 20 with more than 50% vote Democratic versus the 30 with less than 50% did not show statistically significant differences in case rates, though the results were in the same direction – lower case rates among the more Democratic states.

**Figure 3:**
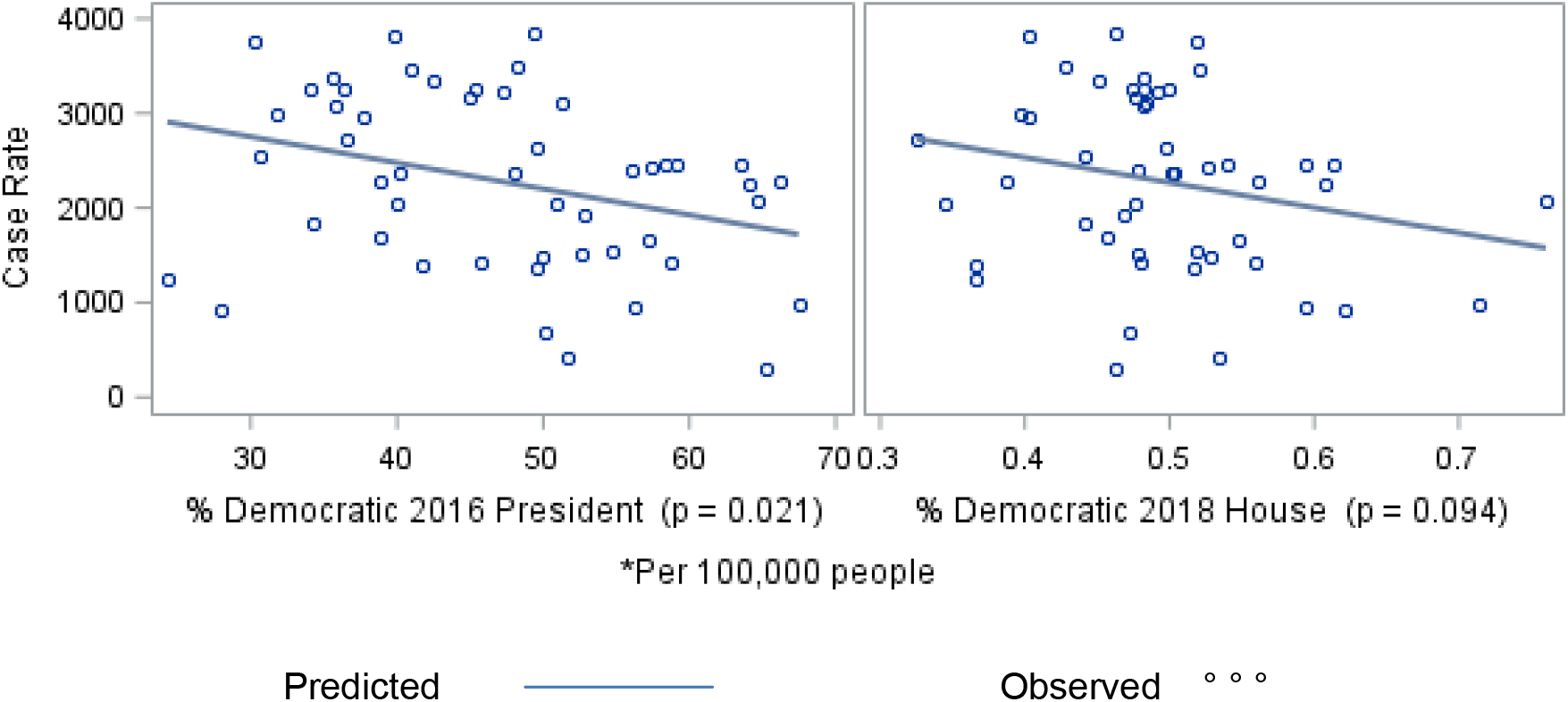
Observed and Predicted Case Rate* versus Percent Democratic in state Vote for 2016 Presidenential Election and for 2018 House Election, in Univariate Analysis

The means of COVID-19 death rates per 100,000 were 49.7 for Republican states, 50.6 for Mixed States, and 69.0 for Democratic states. The differences between the three were not statistically significant (p = 0.320). Of the population characteristics considered, only population density, percentage urbanization, percentage Hispanic, percentage black, and percentage with bachelor’s degree were statistically significantly associated with COVID-19 death rates at the level in univariate analysis. In the multivariate model for death rate only population density and the race/ethnicity variables entered the model as significant, among the state characteristics, and these variables explained 69% of the variation in death rates. Figure 4 plots the results of this model. It stands out that New York, and to a lesser extent Maryland, are notably distant from the diagonal line of equality of observed and predicted death rate.

**FIGURE 4:**
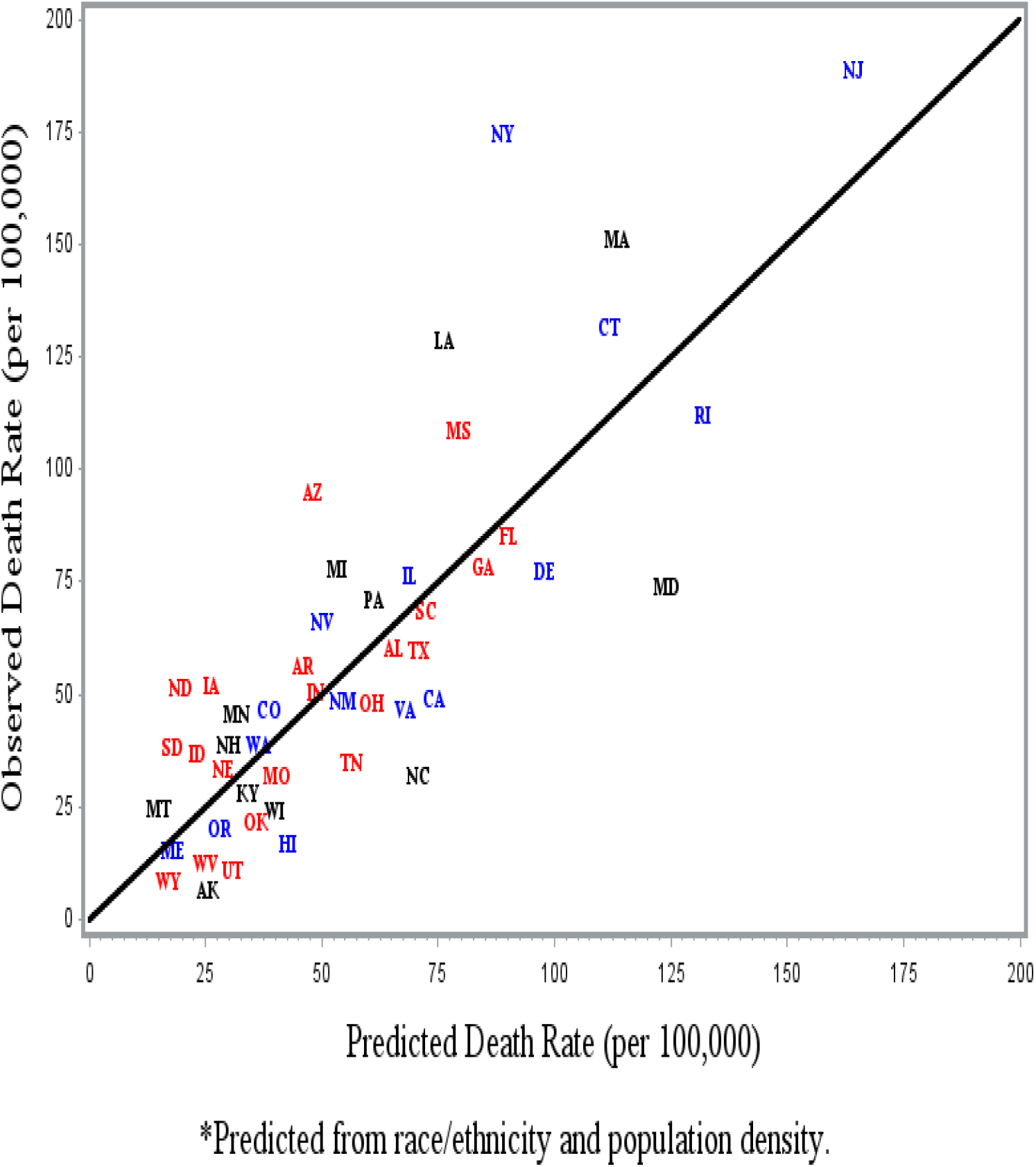
Observed versus Predicted* Cumulative Covid 19 death per 100,000 Population. Party in Power: Democratic=Blue, Republican = Red, Other = Black

When adding the party-in-power variables to this models along with variables which notably affected the differences between the parties, the median of death dates had a major effect on party-in-power differences, and all the state characteristics considered except percentage bachelor’s degree and percentage urbanization had a notable effect on party-in-power differences. The effect of median death date was also statistically significant (p = 0.014), and showed death rate lower by 9 per 100,000 for states with a month later median death date. Adjusting for the demographic, socio-economic, and time factors, Democratic states are estimated to have a 15 per 100,000 COVID-19 lower death rate than Republican states, and Mixed states had a 12 per 100,000 lower death rate than Republican states, but the differences were not statistically significant (Democratic p = 0.194, Mixed p = 0.237). We observed in Figure 4 that New York and Maryland were strong outliers in distance from equality of predicted and observed death rates, so as a sensitivity analysis we fit our final party-in-power death rate model excluding these two states. The results from this were that Democratic states had a 21 per 100,000 lower death rate than Republican-led states (p = 0.032), and mixed government states were lower than Republican states by 7 per 100,000 (p = 0.097), showing the strong effect the two outlying states had on the results.

The adjusted associations between COVID-19 death rate and percentage Democratic vote were not statistically significant for either 2016 presidential vote (p = 0.179) or 2018 House vote (p = 0.964). The results, though not statistically significant, were in the same direction as for party-in-power analysis– the higher the percentage of Democratic vote, the lower the adjusted death rate. Simply diving states into those with more than 50% of the vote Democratic versus less than 50% also did not show a significant difference in death rates, for presidential vote or House vote. However, when dividing states into three groups of the same size, low, middle, and high percentage of Democratic votes, for the 2016 presidential vote the high group had 25 per 100,000 fewer cases than the lower, though not statistically significant (p = 0.096) For the 2018 House election there was little difference between the three groups. When dividing states into three groups of the same size as the party-in-power groups, for the presidential votes the high Democratic percentage group had 37 cases per 100,000 fewer deaths than the lower group (p = 0.023). Using the House votes to form three groups of states with equal size or with party-in-power size found little difference between the groups in COVID-19 death rate.

## Discussion

Independent of the party in power, we have confirmed that various characteristics of U.S. states are associated with case rate for COVID-19 for the period through October 8, 2020. These characteristics include race/ethnicity, median income, median age, percentage of the population aged 19-64 that is uninsured, percentage of the population that is urban, and percentage of the population over the age 25 with a bachelor’s degree (some statistically significant only in the multivariate models). Also independent of the party in power, we have adjusted the rate of COVID-19 in the states, based on the various state characteristics considered, and Republican-led states (those with governor and both chambers of the legislature Republican) more frequently had higher adjusted case rates than Democratic-led states. The regression model adding party-in-power showed that Democratic states had a statistically significantly lower case rate than Republican states, and Mixed government states also had lower case rates, adjusted for the state demographic/socio-economic characteristics. The statistical adjustment reduced the observed, unadjusted differences in case rates by party-in-power. A similar analysis of percentage Democratic vote in either the 2016 presidential election or the 2018 House election, for each election dividing the states into low, middle, and high levels of Democratic votes, and with the groups of the same size as the groups of Republican-led, Mixed, and Democratic-led states, respectively, or with three equally sized groups, found a strong association with case rate for the 2016 presidential election but not for the 2018 House election. Simply dividing the states into two groups according to percentage Democratic vote above 50% or not did not show statistically significant differences between the groups in case rate, for either the presidential or the House vote.

A similar analysis of COVID-19 death rates gave adjusted death rates that were lower in Democratic states and mixed government states than in Republican states, though not statistically significant lower. Whether we found a statistically significant association of death rate with the percentage Democratic of the vote in the 2016 presidential election depended on how we analyzed the percentage, though the associations were in the same direction as those in the party-in-power analysis. The median death date by state for the period of analysis was strongly related to death rate and had a substantial effect in the statistical adjustment of differences by party in power.

It is a task of future research to find reasons for the differences by party in power in COVID-19 case rates. Some of the difference could be explained in part by additional demographic or socio-economic characteristics that are associated with both case rates and party in power. Geographic considerations, such as distance to major airports and the passenger volume at those airports could be important, or spillover effects from an adjacent state. It could be that differences in attitudes of the people in the states explain the lower case rates in Democratic states. There is certainly some suggestion of this in surveys of attitudes towards wearing of masks and social distancing.^21-24^ An additional possible explanation is the difference in public health policies of state governments, such as mask mandates, closing of bars, and support for contact tracing.^25,26^

There are clearly weaknesses in the COVID-19 case data now available. Foremost in the U.S. is the variability among the states in the frequency, timing, and priorities of testing for the virus. There may also be variation in test reliability and in completeness of data collection. We analyze rates of confirmed or probable cases and not the rates of Covid-19 infection. Actual counts of infections will never be available, though estimates of infection rates through antibody testing may be. Rates of hospitalization for COVID-19 would be an alternative disease measure to consider, but 14 states do not offer such data.

The results of our analysis of COVID-19 death rates were inconclusive. There are several potential reasons for this. First, the numbers of deaths are far fewer than those for cases. Second, when enough time has elapsed to ascertain vital status of COVID-19 cases, COVID-19 death rates are a product of case rates times case fatality, where case fatality is the probability of death from COVID-19 among COVID-19 cases. Socio-economic and demographic factors and state public health policies could have different effects on these two components of COVID-19 death rates. Third, there may be additional variation in how “confirmed or probable COVID-19 death” is defined. Fourth, COVID-19 death rates may be changing over time.^27^ Some investigators have found that there is no evidence that age-specific COVID-19 case fatality is changing over time,^28,29^ but changes in the age distribution of cases could nevertheless lead to a change in overall COVID-19 death rates over time. We have tried to adjust for this in our modeling, but it may have been inadequate. Finally, our analysis covers data through October 8, 2020, and possibly includes only half the eventual COVID-19 deaths, so the results of a repeat analysis some months in the future may be more conclusive.

The units of analysis in our study are the 50 U.S. states, not individuals. The choice of the state characteristics considered was driven by studies of individuals, and some of the associations we find are consistent with those found in the study of individuals. Nevertheless, we do not draw conclusions about individuals.

## Data Availability

All data used in this manuscript is publicly available online, include the two listed here and others listed in Data Availability Links.
https://doi.org/10.7910/DVN/IG0UN2
https://doi.org/10.7910/DVN/42MVDX

https://data.census.gov/cedsci/table?q=race%20by%20state&g=0100000US&tid=ACSDP1Y2018.DP05&hidePreview=true

https://github.com/nytimes/Covid-19-data

https://data.census.gov/cedsci/table?q=poverty%20by%20state&t=Income%20and%20Poverty&g=0100000US.04000.001&tid=ACSSPP1Y2019.S0201&hidePreview=true

https://www.census.gov/search-results.html?searchType=web&cssp=SERP&q=median%20household%20income%20by%20state

https://www.census.gov/programs-surveys/geography/guidance/geo-areas/urban-rural/2010-urban-rural.html

https://data.census.gov/cedsci/table?q=race%20by%20state&g=0100000US&tid=ACSDP1Y2018.DP05&hidePreview=true

https://data.census.gov/cedsci/table?q=median%20age%20by%20state&g=0100000US&tid=ACSST1Y2018.S0101&hidePreview=false

https://data.census.gov/cedsci/table?q=ACSST1Y2019.S1501&tid=ACSST1Y2019.S1501&hidePreview=true

https://data.census.gov/cedsci/table?q=uninsured%20by%20state&g=0100000US.04000.001&tid=ACSST1Y2018.S2701&hidePreview=true

https://ballotpedia.org/State_government_trifectas

